# Project CLEAR (ChemicaL Exposure and Awareness Research): A protocol for assessing the availability and chemical composition of skin-lightening products in Northern Manhattan

**DOI:** 10.1101/2025.06.13.25329600

**Authors:** Adana A. M. Llanos, Alexis A. Schaefer, Andrew Turner, Jaia Wingard, Nia Jenkins, Mila Cordero, Jasmine McDonald, Ami R. Zota, Randolph R Singh, Jessica Contreras, Dustin T. Duncan

**Affiliations:** Department of Epidemiology, Mailman School of Public Health, Columbia University Irving Medical Center, New York, NY 10032, USA; Herbert Irving Comprehensive Cancer Center, Columbia University Irving Medical Center, New York, NY, 10032, USA; Department of Biochemistry & Cell Biology, Stony Brook University, 450 Life Sciences Building, Stony Brook, NY 11794, USA; Department of Environmental Health Sciences, Mailman School of Public Health, Columbia University, New York, NY 10032, USA

## Abstract

**Introduction:** Skin lightening products (SLPs) are widely used in communities of color and often contain toxic chemicals such as mercury and hydroquinone, posing serious health risks. Despite regulations, these products remain accessible through illegal sales and deceptive labeling. Targeted marketing in marginalized areas raises environmental justice and public health equity concerns.

**Objectives:** This study employs a novel spatial sampling approach to audit retail stores in Northern Manhattan, assessing the availability of SLPs in relation to neighborhood context. Products will be screened for harmful substances—including per- and polyfluoroalkyl substances (PFAS), parabens, and heavy metals—with results compared to ingredient labels.

**Methods:** Conducted in Northern Manhattan, New York City, this study focuses on neighborhoods with high proportions of Black and Latinx residents. Phase 1 involves a structured audit of 50 retail stores, including beauty supply shops and ethno-cultural retailers. Store selection is guided by spatial sampling and demographic data. Trained research assistants collect detailed information on store environments and individual SLPs. In Phase 2, 20 products will be purchased for laboratory analysis using advanced targeted and non-targeted methods. Analyses will include descriptive statistics, GIS mapping, and comparisons across neighborhoods.

**Results:** We expect beauty supply stores to carry a greater variety and volume of SLPs than ethno-cultural retailers, such as African markets, which are anticipated to sell mostly imported products. Chemical testing is expected to show that a substantial portion of SLPs contain hazardous chemicals, including some not disclosed on product labels.

**Conclusions:** Project CLEAR combines spatial methods and laboratory science to map SLP availability and assess chemical risks in Northern Manhattan. By linking store-level data with neighborhood demographics, the study highlights structural inequities and environmental racism. Findings will support future research, inform policy and regulatory efforts, and strengthen community advocacy for safer, transparent skincare products.

## Introduction

Skin lightening products (SLPs) represent a rapidly growing sector of the global beauty market, projected to reach $15.7 billion by 2030 [1]. SLPs – cosmetics including creams, gels, lotions and soaps that are used to chemically lighten the skin for aesthetic reasons – are frequently used among individuals with darker shades of skin to achieve a lighter complexion. Although data is limited, research has indicated disproportionate use among racial and ethnic minorities and immigrants, driven by Eurocentric beauty standards rooted in racism, colorism, and colonialism [2, 3]. This practice exemplifies environmental racism, as marginalized communities face targeted marketing of toxic products. Evidence overwhelmingly shows Asian, Latinx, and Black women face heightened risks of adverse health effects due to lifelong SLP use, product use early in childhood, and sustained use over critical windows of susceptibility [4, 5]. Thus, addressing this public health problem can help advance beauty justice—an emerging area of research that frames racial and ethnic inequalities in exposure to potentially toxic chemicals in PCPs as a matter of environmental justice, and advocates for equitable access to safer products as a strategy to reduce exposure and related health inequities [6, 7].

Common chemicals of concern (CoCs) used in skin bleaching products include mercury, lead, hydroquinone, hydrogen peroxide, corticosteroids, polyhydroxy acids, and azelaic acid. These substances are known to have adverse effects on health [8-10]. Hydroquinone, which has been banned for use in over the counter (OTC) products by many countries, including the United States through legislation that classified hydroquinone as a Category II drug as part of the CARES Act of 2020 [11] with regulation expanded as part of the Modernization of Cosmetics Regulation Act (MoCRA) of 2022 [12], has been associated with many harmful health effects ranging from acute conditions like irritant contact dermatitis to cancer [13]. Emerging evidence also shows that per- and polyfluoroalkyl substances (PFAS) are included in SLP formulations to alter the product texture [14-17]. Known dermatological effects associated with these CoCs include skin atrophy, ochronosis, scarring, thinning, breaking, and infections, all of which can cause keloidal scarring, perioral dermatitis, contact allergic and irritant dermatitis, acneiform eruptions, striae, hypertrichosis, and telangiectasias [18]. Common infections associated with CoCs in SLPs include both fungal infections (*Candida*, dermatophyte) and bacterial infections (pyoderma, folliculitis, furuncles, impetiginous lesions, erysipelas) [18]. These chemicals are also associated with increased liver and kidney toxicity, proteinuria, and neurotoxic effects [19]. In severe cases, exposure to the CoC clobetasol propionate, a corticosteroid, can cause skin thinning, hormonal imbalances, and other systemic effects. Clobetasol propionate reduces inflammation, which in turn suppresses melanin production, resulting in lighter skin; as a result, it is commonly used in SLPs. The presence of this chemical in OTC products is particularly concerning given its potency and interaction with the central nervous system and essential organs [20, 21].

Mercury, a CoC often found in SLPs – although United States Food and Drug Administration regulations prohibit its use in most cosmetics. Specifically, inorganic mercury salts are frequently added intentionally to SLPs to inhibit melanin production in the skin [19]. In addition to dermal exposure from topical use of SLPs, individuals, including children, may also be exposed to mercury through inhalation of mercury vapors emitted by products, highlighting both personal and household exposure risks [22-24]. A systematic review by Bastinansz, et al. highlights the dangers of large-scale mercury use in the SLP market [19]. An evaluation of 41 peer reviewed scientific literature published between 2000 and 2022 from 22 countries, the investigators estimated an overall pooled central median mercury level of 0.49μg/g; (interquartile range [IQR]: 0.02–5.9) from 787 skin-lightening products [19]. Using human biomarker measurements from 863 individuals, analysis also showed that those with high levels of total mercury in urine, blood, and hair presented with signs of mercury intoxication, nephrotic syndrome, membranous glomerulonephritis, and hypertension [19]. These findings reiterate that mercury is commonly found in SLPs across numerous countries, exposing users around the world to relatively high levels of mercury through personal product use and household inhalation.

Evidence from epidemiological, clinical, and environmental toxicology studies published in the last two decades strongly link mercury exposure and carcinogenesis, particularly through the disruption of estrogen receptor ERK1/2, JNK, NADPH-oxidase, and Nrf2 signaling cellular signaling pathways [25]. This disturbance mechanism interrupts highly regulated cell growth, proliferation, and stress responses, playing a significant role in cancer pathogenesis due to reactive oxygen species (ROS) overproduction and subsequent oxidative DNA damage [19, 25-27]. Despite the federal ban on many of these products due to hazardous chemical ingredients, SLP use has not deterred [4, 14, 20, 28].

Skin lightening has long been documented in African, Asian and Caribbean countries; however, the use of SLPs is increasingly on the rise in the United States (US), especially among immigrant populations [29]. SLP use is recognized by the World Health Organization (WHO) as a significant global public health and environmental health issue. A New York City (NYC)-based study found 25% of Northern Manhattan residents reported SLP use, with higher rates among Asian women and immigrants [29]. The NYC Department of Health and Mental Hygiene (NYC DOHMH) recently developed “The Intervention Model for Contaminated Consumer Products”, which includes surveillance and enforcement of mercury-containing SLPs and also risk communication [30]. Their research has indicated that some SLPs reach up to 40,000 times the permitted limit of mercury [30]. To increase awareness of this, the NYC DOHMH created a list of specific SLPs to avoid, categorized by country of origin, with a majority coming from Pakistan [31]. On December 23, 2022, Governor Hochul signed into law a bill that prohibits the sale of any cosmetic product containing mercury. Although there are some exceptions to this law (e.g., trace amounts of mercury that may be unavoidable under conditions of good manufacturing practices or necessary use of mercury as a preservative), this ban, which took effect on June 1, 2023, increases public awareness regarding the potential harms of SLP use.

While these bans have increased awareness about CoCs in SLPs, they have also contributed to the illegal sale and rebranding of products, often with harmful ingredients omitted from labels [10]. Despite bans, products remain widely available due to illegal rebranding and label fraud [15-17, 32]. This deceptive marketing has raised significant alarm, particularly in NYC where cultural beauty standards and widespread availability encourage product use. Due to insufficient regulation and lack of transparency in the ingredients lists of SLPs, individuals throughout NYC may unknowingly expose themselves to dangerous levels of toxic substances like mercury and hydroquinone, posing serious public health risks [4, 33]. This deception also makes research on the impacts of SLP use particularly difficult and needed [4, 14], especially post-2022.

To address critical gaps in understanding the unequitable distribution of SLPs in Northern Manhattan, this project investigates the targeted marketing of hazardous SLPs in vulnerable communities, where products often evade regulatory oversight [30, 31]. Preliminary audits reveal stark disparities in neighborhoods like East Harlem where African and Caribbean bodegas prominently stock imported SLPs containing banned substances (e.g., mercury-laced creams from Côte d’Ivoire), compared to retailers in affluent NYC zip codes where an absence of SLPs has been documented [34]. Based on this research, we seek to further explore the landscape of SLPs in the consumer market in Northern Manhattan, using spatial sampling and laboratory science methods.

## Project Objectives

### Overview

This pilot study—incorporating principles of the Health Equity Research Production Model [35]—employs an innovative methodological approach to systematically characterize the landscape of SLP availability in neighborhood retail stores across Northern Manhattan, with a specific focus on assessing variation in product availability by neighborhood context. Utilizing a rigorous spatial sampling protocol, the study will conduct a neighborhood audit of 50 retail stores to comprehensively evaluate the presence and distribution of SLPs within the local consumer market Northern Manhattan. In the second phase, selected products will undergo screening and quantification of CoCs to determine the prevalence of potentially harmful ingredients in products available for purchase. Store census data will be matched with demographic and socioeconomic indicators from the American Community Survey, including neighborhood racial and ethnic composition and the neighborhood deprivation index. Approximately 20 products identified through this audit will be carefully selected and purchased for laboratory analysis to quantify the presence of CoCs, including phthalates, parabens, heavy metals, and PFAS—toxicants to which individuals using SLPs may be exposed. Employing non-targeted analysis will allow for the detection of undisclosed chemicals in SLPs and the comparison of results with product ingredient lists. This study was reviewed by the Columbia University Institutional Review Board (IRB) and deemed not to involve human subjects research, as it does not include consenting individuals or soliciting their participation in any study-related activities.

### Specific Aims and Hypotheses

The Specific Aims are: (1) to survey the landscape of SLP availability (e.g., number of products, types, pricing, label details, and listed ingredients) in Northern Manhattan; and (2) to purchase and analyze up to 20 SLPs available in the local market to quantify the presence of harmful chemicals commonly found in these products. Based on our environmental justice framework, for Aim 1, we anticipate a wide variety of SLPs in the local consumer market, representing multiple brands and product types, with many manufactured outside the United States. In Aim 2, we expect to detect CoCs, including phthalates and parabens (endocrine-disrupting compounds), mercury, cadmium, and arsenic (heavy metals), as well as short-chain perfluoroalkyl acids (e.g., perfluorobutane sulfonic acid [PFBS], perfluorobutanoic acid [PFBA], and perfluoropentanoic acid [PFPeA]), perfluorohexanesulfonic acid (PFHxS), and their precursors in SLPs available in Northern Manhattan. Additionally, we hypothesize that some substances will be present at elevated concentrations and that many will not be disclosed on product labels.

## Methods

### Setting

This study will take place in New York, NY with a specific focus on the Northern Manhattan region **(Figure 1)**. New York City is home to approximately 7.9 million residents and spans roughly 300 square miles. Manhattan, one of the city’s five boroughs, has an estimated population of 1.6 million within a compact area of about 22.7 square miles. Northern Manhattan generally includes the neighborhoods of Harlem, Washington Heights, and Inwood—communities with rich cultural histories and diverse populations. Harlem is home to around 197,000 residents, while Washington Heights and Inwood have estimated populations of 172,800 and 38,400, respectively. These neighborhoods provide a critical context for examining environmental health disparities and access to consumer products.

**Figure 1.**
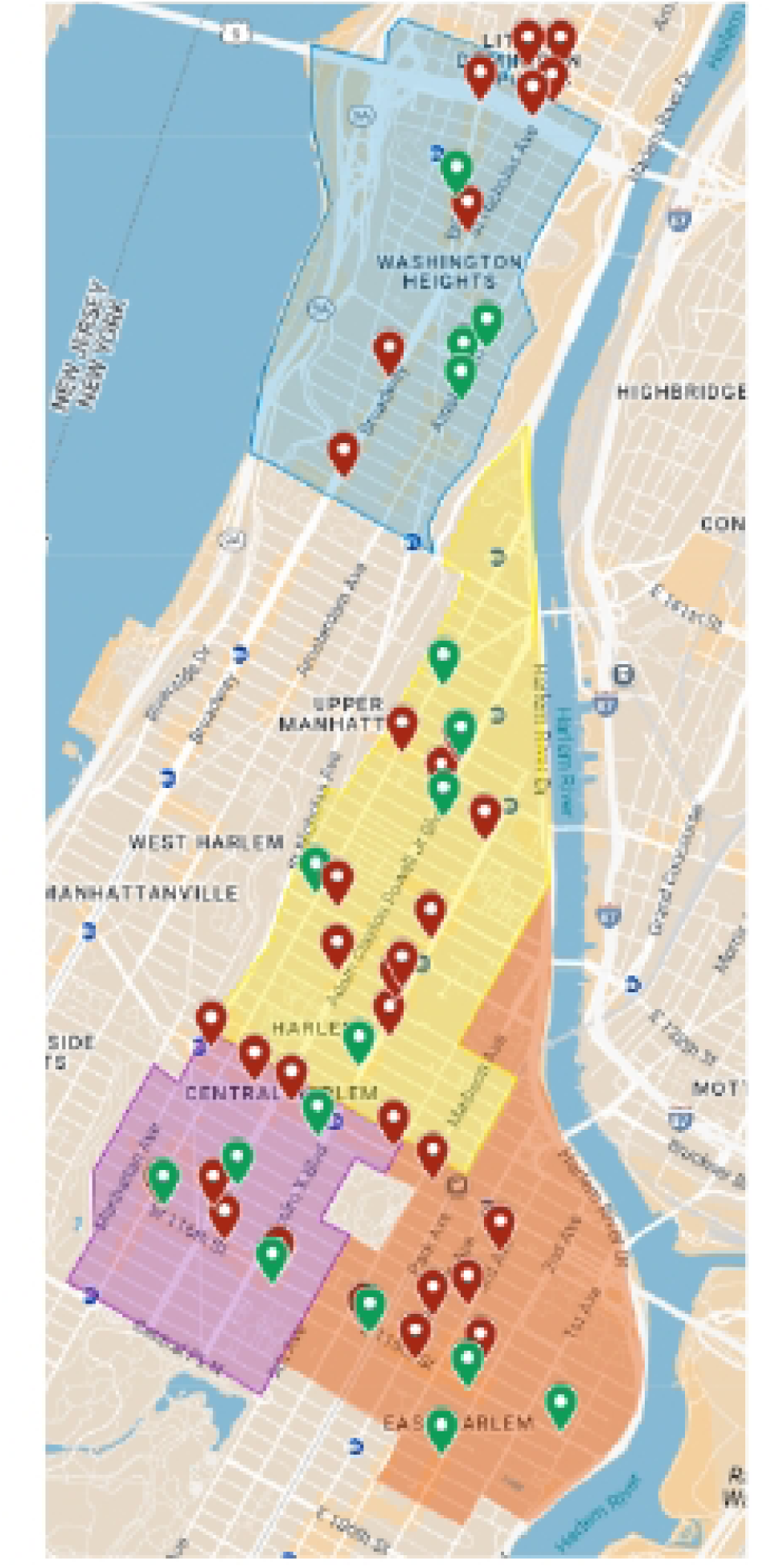
Map of Target Retail Stores Selected for Field Research in Northern Manhattan: Colored regions represent neighborhood boundaries: Washington Heights (blue), Central Harlem North (yellow), Central Harlem South (purple), and East Harlem (orange). Pins indicate selected stores, with green markers representing ethno-cultural stores and red markers representing beauty supply stores. Map created using Google Maps (© Google, 2025).

### Phase 1

To accomplish Aim 1, we will systematically assess SLP availability across Northern Manhattan through a survey of 50 stores in specifically selected neighborhoods **(Figure 1**, **Table 1)**. Our neighborhood selection strategy will employ a demographic-driven approach using defined Neighborhood Tabulation Areas (NTAs) from the NYC Department of City Planning. Selection criteria will prioritize Northern Manhattan areas with >50% Black and Latinx residents, as these populations have been identified in previous research as target consumers for SLPs [28, 34]. Based on these criteria, the study will focus on four selected NTAs—Washington Heights South, Central Harlem North, Central Harlem South, and East Harlem—which together provide geographic diversity across Northern Manhattan while maintaining proximity to our institution’s catchment area (Columbia University). Neighborhood demographic profiles, including racial and ethnic composition and socioeconomic indicators (median household income, educational attainment, and neighborhood deprivation index), will be obtained from the American Community Survey 5-year estimates (2017-2021) to contextualize our findings. This neighborhood selection approach allows us to examine potential differences in SLP availability and marketing across demographically similar yet culturally distinct communities in Northern Manhattan.

**Table 1.** List of Stores to Visit During Field Research.

| Neighborhood | Store Name | Store Type | Store Code |
| --- | --- | --- | --- |
| Washington Heights South | Feel Beauty Supply | Beauty Supply | WSH-FBS |
| Washington Heights South | Total Beauty Supply | Beauty Supply | WSH-TBS |
| Washington Heights South | La Bella Beauty Supplies | Beauty Supply | WSH-LBB |
| Washington Heights South | In Beauty Supply | Beauty Supply | WSH-IBS |
| Washington Heights South | Bonita Beauty Supply | Beauty Supply | WSH-BBS |
| Washington Heights South | Young Hee Beauty Supply Store | Beauty Supply | WSH-YHB |
| Washington Heights South | Natanael Beauty Supply Inc | Beauty Supply | WSH-NBS |
| Washington Heights South | Sara Health & Beauty Inc | Beauty Supply | WSH-SHB |
| Washington Heights South | Señor Santo Entierro de Xalpathlahuac | Ethno-Cultural | WSH-SSE |
| Washington Heights South | Lindo Amanecer Grocery Corp | Ethno-Cultural | WSH-LAG |
| Washington Heights South | 2143 El Patron Mini Market Corp | Ethno-Cultural | WSH-EPM |
| Washington Heights South | Las Palmeras Deli Grocery | Ethno-Cultural | WSH-LPD |
| Central Harlem North | Diallo Beauty Supply Inc | Beauty Supply | CHN-DBS |
| Central Harlem North | Red Envy Hair Care | Beauty Supply | CHN-REH |
| Central Harlem North | Missy Hair Boutique New York | Beauty Supply | CHN-MHB |
| Central Harlem North | Zamzam Health & Beauty Supply | Beauty Supply | CHN-ZHB |
| Central Harlem North | Lenox Beauty Supply | Beauty Supply | CHN-LBS |
| Central Harlem North | Mama Aicha Beauty Supply | Beauty Supply | CHN-MAB |
| Central Harlem North | Dominican Star Beautys Salon | Beauty Supply | CHN-DSB |
| Central Harlem North | Cosmetic Beauty Supply Inc | Beauty Supply | CHN-CBS |
| Central Harlem North | Today Beauty Supply | Beauty Supply | CHN-TBS |
| Central Harlem North | Sugg's World Beauty Supply | Beauty Supply | CHN-SWB |
| Central Harlem North | Beauty R Us | Beauty Supply | CHN-BRU |
| Central Harlem North | Garcia's Tulcingo Mexican Products | Ethno-Cultural | CHN-GTM |
| Central Harlem North | Bahini African Market | Ethno-Cultural | CHN-BAM |
| Central Harlem North | Farida Halal African Market | Ethno-Cultural | CHN-FHA |
| Central Harlem North | Dalaba African American and Caribbean Market Inc | Ethno-Cultural | CHN-DAA |
| Central Harlem North | African Market | Ethno-Cultural | CHN-AMK |
| Central Harlem South | Dabakh Boutique Solutions | Beauty Supply | CHS-DBS |
| Central Harlem South | Belle Nubian Skin Care | Beauty Supply | CHS-BNS |
| Central Harlem South | Apollo Beauty Supply | Beauty Supply | CHS-ABS |
| Central Harlem South | Apollo Beauty Land | Beauty Supply | CHS-ABL |
| Central Harlem South | Orange Beauty Supply | Beauty Supply | CHS-OBS |
| Central Harlem South | Max Beauty | Beauty Supply | CHS-MBY |
| Central Harlem South | Beauty Sensation | Beauty Supply | CHS-BSN |
| Central Harlem South | Khady Beauty Supply | Beauty Supply | CHS-KBS |
| Central Harlem South | Touba African Clothing | Ethno-Cultural | CHS-TAC |
| Central Harlem South | Maliba African Market | Ethno-Cultural | CHS-MAM |
| Central Harlem South | Adja Khady | Ethno-Cultural | CHS-AKY |
| Central Harlem South | Malcolm Shabazz Harlem Market | Ethno-Cultural | CHS-MSH |
| East Harlem | Luxurhair Collection | Beauty Supply | EAH-LCN |
| East Harlem | Real Beauty Center | Beauty Supply | EAH-RBC |
| East Harlem | Fajas Luxurys Manhattan | Beauty Supply | EAH-FLM |
| East Harlem | Total 125 Beauty Supply | Beauty Supply | EAH-TBT |
| East Harlem | Beauty Supply of New York | Beauty Supply | EAH-BSO |
| East Harlem | Dubai Cosmetics | Beauty Supply | EAH-DCS |
| East Harlem | Total Beauty Supply | Beauty Supply | EAH-TBS |
| East Harlem | La Costeñita Grocery | Ethno-Cultural | EAH-LCG |
| East Harlem | El Pueblo Mexicano Grocery | Ethno-Cultural | EAH-EPM |
| East Harlem | Touba African Market Dress Flavor | Ethno-Cultural | EAH-TAM |
| East Harlem | Fifi Market | Ethno-Cultural | EAH-FMK |

We will develop a two-tiered store classification system to systematically identify retail locations likely to sell SLPs. Primary sources, our priority for data collection, will include beauty supply stores and ethno-cultural retailers. In this study, we define ethno-cultural retailers as establishments specifically marketing imported products or merchandise catering to specific ethnic or immigrant communities (e.g., Mexican product suppliers, specialty African cosmetic retailers). Secondary sources will consist of boutique pharmacies which we define as independently owned, non-chain pharmacies. These secondary sources will serve as contingency sampling locations and will be included only if the number of primary stores is insufficient to achieve our target sample size of 50 establishments. This classification approach ensures focused data collection at locations with the highest likelihood of SLP availability while maintaining flexibility to reach our sampling goals.

We will employ a systematic digital mapping approach to identify stores likely to sell SLPs within our selected neighborhoods. Using Google Maps as our primary search tool, we will conduct targeted searches using standardized terminology. For beauty supply stores, search terms will include “beauty supply,” “hair and beauty,” beauty wholesale” while ethno-cultural stores will be identified using terms like “African markets”, “Caribbean markets”, and neighborhood-specific ethnic group descriptors relevant to each NTA’s demographic composition. Boutique pharmacies will be located using search terms “pharmacy” and “drug store,” followed by manual filtering to exclude chain establishments. Neighborhood boundaries will follow official NYC Department of City Planning NTA delineations, with a 2-block extension in border areas to reflect realistic shopping patterns. This extension will be particularly important for commercial corridors split by administrative boundaries, such as 181st Street in Washington Heights South, where the official boundary bisects a popular shopping area. All identified stores will be cataloged in Google Maps lists and duplicated in MyMaps to create comprehensive neighborhood-specific store databases with precise geolocation data for field data collection.

Our sampling strategy aims to comprehensively capture SLP availability across Northern Manhattan using a census approach of primary stores. Preliminary mapping identified 51 primary stores (34 beauty supply stores and 17 ethno-cultural retailers) across the four selected neighborhoods **(Figure 1)**. If this number remains stable during implementation, we will conduct a complete census of all primary stores, maximizing our understanding of the SLP market landscape. If our preliminary count changes substantially and exceeds our capacity, we will implement a stratified random sampling approach proportional to the number of stores in each NTA to maintain our target sample size of 50 establishments. Conversely, if any primary stores are found to be permanently closed or inaccessible during data collection, we will supplement the sample with strategically selected secondary stores (boutique pharmacies) to maintain our minimum sample size. This sample size is sufficient for a pilot study, as it will capture most specialized retailers likely to carry SLPs while remaining logistically feasible within our timeline. Overall, the comprehensive sampling approach will provide a robust assessment of SLP availability in the specialized retail sector across Northern Manhattan.

Data collection will begin with a pilot phase in Washington Heights South, selected for its proximity to Columbia University to facilitate oversight and procedural refinement. During this phase, Research Assistants (RAs) will undergo comprehensive training, covering store engagement protocols, safety procedures, and cultural sensitivity training, including role-playing exercises to prepare for potential in-store scenarios. Our research team includes bilingual RAs fluent in Spanish, and our team composition reflects the diverse demographic characteristics of the populations residing in our target neighborhoods. RAs will work in pairs to systematically document store-level and product-specific information using standardized Qualtrics survey forms on smartphones. The pairing system ensures safety, improves data quality through dual verification, and provides support in addressing potential language barriers if needed.

At the product level, data collection will capture brand name, product type (cream, serum, mask, or liquid spray), price, and complete photographic documentation of packaging, ingredients list, and product identifiers **(Table 2)**. Store-level data collection will assess the broader retail context, including neighborhood environment, exterior and interior advertisements, product placement, pricing strategies, stock levels, and target demographic indicators represented in marketing materials **(Table 2)**. This dual-level measurement strategy will enable analysis of both individual SLPs and the retail ecosystems in which they are marketed. Regular debriefing sessions will be conducted to address emerging challenges, ensure protocol consistency across neighborhoods, and refine data collection procedures as needed.

**Table 2.**
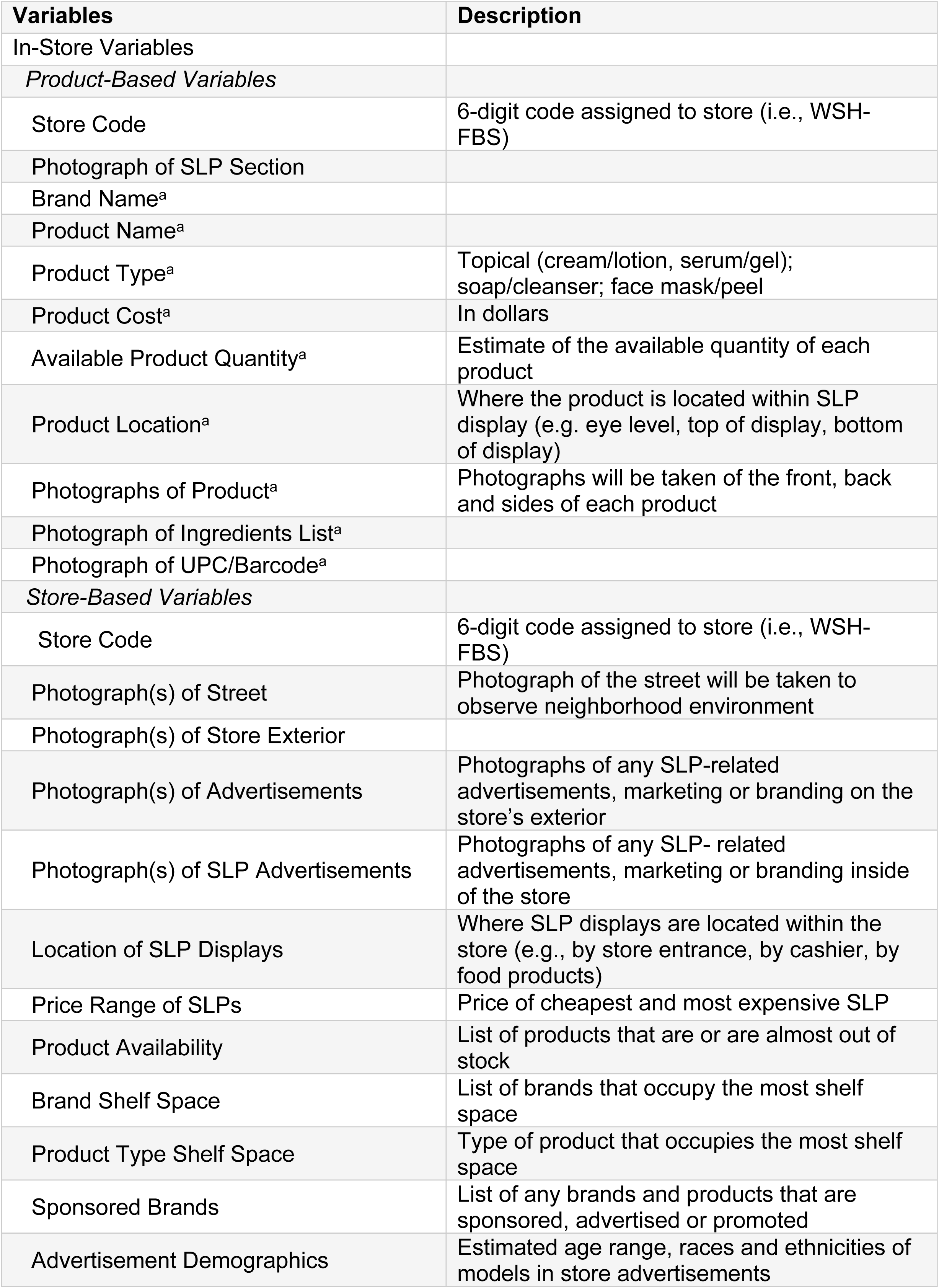
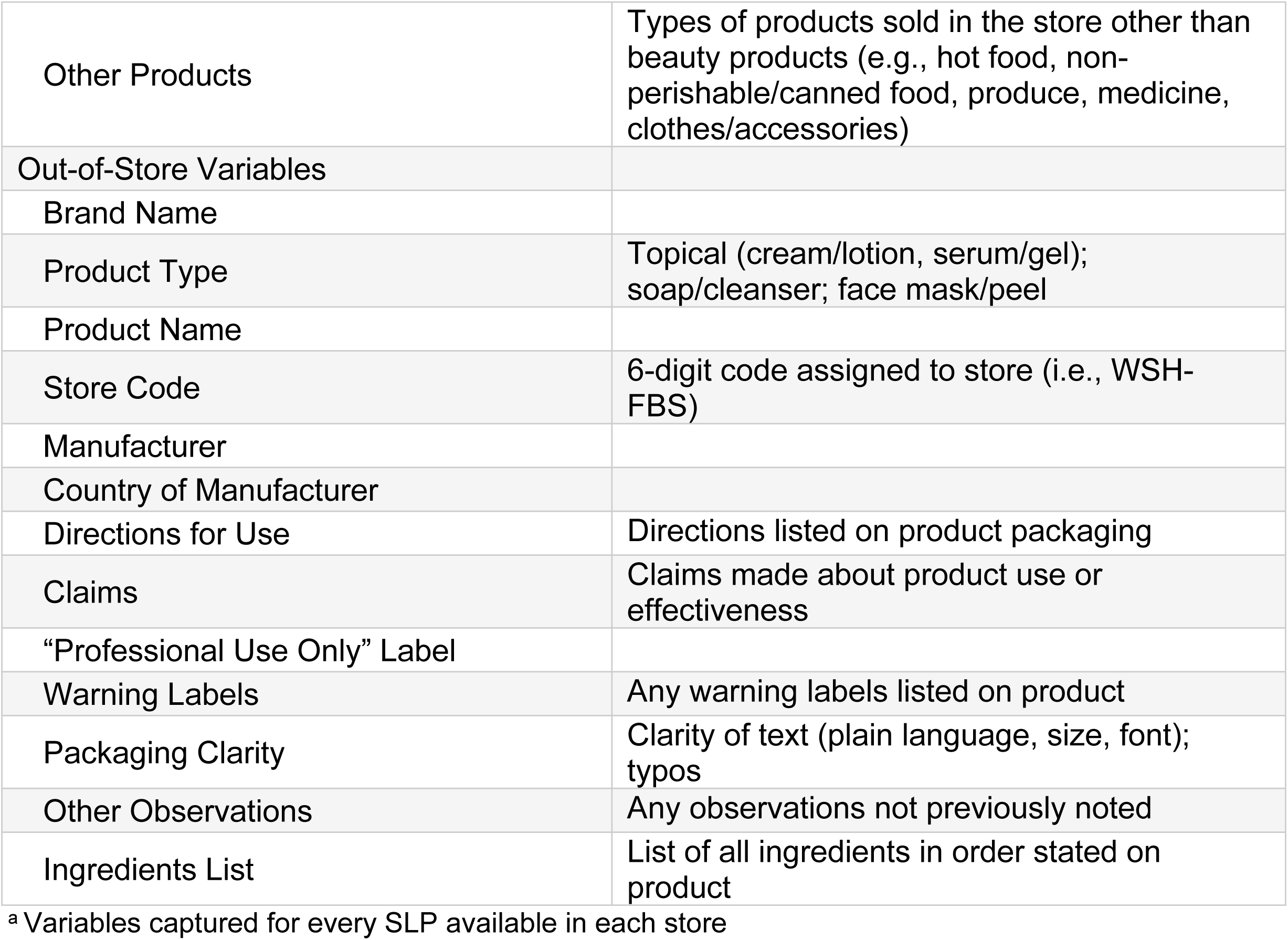
Variables to Collect in and Out of Store During Field Research.

| **Table 2. Variables to Collect in and Out of Store During Field Research** |  |
| --- | --- |
| **Variables** | **Description** |
| In-Store Variables |  |
| *Product-Based Variables* |  |
| Store Code | 6-digit code assigned to store (i.e., WSH-FBS) |
| Photograph of SLP Section |  |
| Brand Name<sup>a</sup> |  |
| Product Name<sup>a</sup> |  |
| Product Type<sup>a</sup> | Topical (cream/lotion, serum/gel); soap/cleanser; face mask/peel |
| Product Cost<sup>a</sup> | In dollars |
| Available Product Quantity<sup>a</sup> | Estimate of the available quantity of each product |
| Product Location<sup>a</sup> | Where the product is located within SLP display (e.g. eye level, top of display, bottom of display) |
| Photographs of Product<sup>a</sup> | Photographs will be taken of the front, back and sides of each product |
| Photograph of Ingredients List<sup>a</sup> |  |
| Photograph of UPC/Barcode<sup>a</sup> |  |
| *Store-Based Variables* |  |
| Store Code | 6-digit code assigned to store (i.e., WSH-FBS) |
| Photograph(s) of Street | Photograph of the street will be taken to observe neighborhood environment |
| Photograph(s) of Store Exterior |  |
| Photograph(s) of Advertisements | Photographs of any SLP-related advertisements, marketing or branding on the store's exterior |
| Photograph(s) of SLP Advertisements | Photographs of any SLP- related advertisements, marketing or branding inside of the store |
| Location of SLP Displays | Where SLP displays are located within the store (e.g., by store entrance, by cashier, by food products) |
| Price Range of SLPs | Price of cheapest and most expensive SLP |
| Product Availability | List of products that are or are almost out of stock |
| Brand Shelf Space | List of brands that occupy the most shelf space |
| Product Type Shelf Space | Type of product that occupies the most shelf space |
| Sponsored Brands | List of any brands and products that are sponsored, advertised or promoted |
| Advertisement Demographics | Estimated age range, races and ethnicities of models in store advertisements |
| Other Products | Types of products sold in the store other than beauty products (e.g., hot food, non-perishable/canned food, produce, medicine, clothes/accessories) |
| Out-of-Store Variables |  |
| Brand Name |  |
| Product Type | Topical (cream/lotion, serum/gel); soap/cleanser; face mask/peel |
| Product Name |  |
| Store Code | 6-digit code assigned to store (i.e., WSH-FBS) |
| Manufacturer |  |
| Country of Manufacturer |  |
| Directions for Use | Directions listed on product packaging |
| Claims | Claims made about product use or effectiveness |
| “Professional Use Only” Label |  |
| Warning Labels | Any warning labels listed on product |
| Packaging Clarity | Clarity of text (plain language, size, font); typos |
| Other Observations | Any observations not previously noted |
| Ingredients List | List of all ingredients in order stated on product |

<sup>a</sup> Variables captured for every SLP available in each store

Given that our research involves observational data collection in commercial establishments, we will implement several ethical safeguards. All observations will be conducted during regular business hours without disrupting store operations. RAs will adhere to standardized interaction protocols when engaging with store employees, including prepared responses to common inquiries. No personally identifiable information about store employees or customers will be collected. Photography will be restricted to products and displays, avoiding the capture of individuals. RA safety will be prioritized through a buddy system, regular check-ins, and comprehensive training on navigating potentially uncomfortable interactions.

### Phase 2

To accomplish Aim 2, we will purchase a range of SLPs from the 50 store locations sampled in Aim 1. Our goal is to characterize the SLPs currently available by selecting products identified during the neighborhood survey. Although the exact number of brands and products available is unknown, we will purchase each type of SLP encountered (e.g., creams, soaps). Priority will be given to products that are most widely available, particularly those listing ingredients known to pose health risks (e.g., hydroquinone, mercury). Additionally, products of potential concern based on a preliminary review of ingredient labels will be incorporated. We will first select the 15 most commonly available products, then supplement with five additional products based on either ingredient concerns or to ensure representation of all SLP types (e.g., SLP powders). This selection strategy balances relevance and feasibility, ensuring comprehensive product characterization for analysis.

Up to 20 SLPs will be screened for CoCs using two-dimensional gas chromatography coupled with mass spectrometry (GCxGC-MS) through non-targeted analysis and retention time-only standards. The contract lab maintains readily available standard mixtures, including a paraben mixture (methylparaben, ethylparaben, n-propylparaben, iso-propylparaben, n-butylparaben, isobutylparaben, and benzylparaben), a phthalate mixture (benzyl butyl phthalate, bis(2-ethylhexyl) adipate, bis(2-ethylhexyl) phthalate, dibutyl phthalate, diethyl phthalate, dimethyl phthalate, and di-n-octyl phthalate), and an Environmental Protection Agency (EPA) 8270 mixture containing various semi volatile environmental chemicals of interest. Results will be generated using automated processing systems, providing estimated concentrations of each detected chemical per sample. Chemical identifications, except for those with established retention time standards, will rely on NIST library matching and will be considered tentative. Products will also be screened for metals using inductively coupled plasma mass spectrometry (ICP-MS). Results will be delivered electronically in a spreadsheet containing, at a minimum, the concentration of each target chemical per sample, sample ID, dates/times of receipt and analysis. Finally, ingredient lists from SLP product labels will be cross-referenced with chemical screening results to characterize available products and assess label accuracy.

### Training for Phases 1 and 2

In preparation for Phases 1 and 2, a comprehensive training was conducted in April 2025 with the research team, including the Llanos Lab and the Columbia Spatial Epidemiology Lab. The training encompassed a detailed review of the protocol, an examination of a published study employing systematic social observation [36, 37], the study timeline and instruction on the standardized protocol for capturing SLP information. In May 2025, four research assistants, working in pairs of one English and one Spanish speaker, conducted initial field observations to pilot test the study procedures. During this phase, eight stores total were visited between the two pairs, with two stores surveyed in each target neighborhood. At the end of May 2025, the team held a debriefing session to review field experiences and discuss preliminary findings. Insights from this iterative process led to key refinements in the data collection protocol, including updates to the Qualtrics tools to improve efficiency and streamline the process.

### Planned Analyses

#### Spatial Distribution Analysis

Store audit data will be analyzed using descriptive statistics to characterize the SLP landscape. We will calculate frequencies and proportions of SLP availability by product type, price range, and stated ingredients across neighborhoods. Geographic information system (GIS) mapping will be employed to visualize the spatial distribution of SLPs in relation to sociodemographic characteristics obtained from American Community Survey data. This spatial analytic approach will facilitate the identification of potential "hotspots" of SLP availability and enable an assessment of whether product types exhibit clustering within specific community contexts.

#### Neighborhood Comparative Analysis

We will examine associations between neighborhood characteristics (e.g., racial and ethnic composition, socioeconomic indicators) and patterns of SLP availability. Proportions of stores carrying SLPs will be calculated for each neighborhood, and product characteristics will be compared across neighborhoods using chi-square tests where appropriate. This analysis will help determine whether environmental injustice observed in other consumer product categories extends to the SLP market in Northern Manhattan [29, 38].

#### Chemical Composition Analysis

For chemical analyses, concentration of each CoC identified in the products will be reported as ranges across the sampled products. Comparisons between labeled ingredients and detected chemicals will be conducted to assess transparency in product labeling. Products will be categorized by chemical profile, price point, and target market to identify patterns associated with harmful chemical exposures from SLPs in Northern Manhattan communities. These results will inform future research on exposure assessment and risk characterization, support the development of educational materials for consumers and healthcare providers, and contribute to policy discussions regarding the regulation of harmful ingredients in personal care products.

### Timeline

Given the one-year duration of the pilot project, a structured and feasible timeline was developed to ensure the successful achievement of all objectives. The initial phase will focus on preparatory activities, including staff training, the development of research protocols and instruments, and obtaining Institutional Review Board (IRB) approval. The second phase will involve the implementation of neighborhood-based survey research. During this period, research staff will identify and recruit participating retail locations. Once the store list is finalized, in-store observations will be conducted to document the availability and characteristics of SLPs. A selected sample of SLPs will be purchased for further analysis during this time. The third phase will consist of laboratory testing of the acquired products, comprehensive data analysis, and preparation of abstracts and manuscripts for data dissemination.

## Results – Expected Findings

### Store Type and SLP Availability Patterns

We anticipate significant variation in SLP availability by store type within our two-tiered classification system. Beauty supply stores are expected to have both higher prevalence and a greater diversity of SLPs compared to ethno-cultural retailers, given their exclusive focus on appearance-altering products. Preliminary observations suggest that beauty supply stores may dedicate entire sections to SLPs, potentially offering five to ten distinct products per location.

Among ethno-cultural retailers, we anticipate considerable heterogeneity in the availability of SLP, with African markets expected to exhibit the highest prevalence of such offerings. This expectation is supported by prior research documenting high SLP usage rates in West African countries, where prevalence ranges from 25% in Mali to 77% in Nigeria [39]. Given that neighborhoods like Harlem have significant West African immigrant populations, we expect to find a greater availability of imported SLPs in these areas. In contrast, ethno-cultural retailers serving Central American immigrant communities may have more variable SLP availability.

We also anticipate that the chemical composition of products will vary by store type, with beauty supply stores likely carrying more products whose ingredient lists comply with US regulations, while ethno-cultural retailers may stock a greater proportion of imported products containing higher levels of restricted ingredients such as mercury, hydroquinone, and corticosteroids. This anticipated pattern aligns with our environmental justice approach, as products with potentially higher toxicity levels may be more accessible in immigrant-dense neighborhoods.

### Product Origin and Regulatory Compliance

Given the global nature of the SLP market, we expect many products available in Northern Manhattan stores to be manufactured outside the US. Products from Pakistan, India, Mexico, the Dominican Republic, and various African nations are likely to be especially prominent in ethno-cultural retailers. We anticipate that a substantial proportion of these imported products may lack complete English-language ingredient lists or fail to comply with FDA labeling requirements, potentially limiting consumers’ ability to make informed decisions about associated health risks.

### Chemical Analysis Findings

For Aim 2, we expect chemical analysis to detect multiple CoCs in the selected products. Based on the 2023 Zero Mercury Working Group (ZMWG) report, which detected mercury above legal limits in 90% of tested SLPs, and analysis conducted by Bastinansz et al., we anticipate that at least 60-70% of tested products will contain mercury levels exceeding the legal threshold of 1 ppm, with concentrations potentially ranging from 1 ppm to over 75,000 ppm [19, 27]. We also expect to identify phthalates, parabens, and PFAS compounds, many of which may not be disclosed on product labels.

## Conclusions

Project CLEAR advances the field by introducing a novel, integrative approach to evaluating the consumer environment and chemical exposures associated with SLPs. Through a spatially rigorous survey of SLP availability across neighborhood stores in Northern Manhattan, coupled with targeted and non-targeted laboratory analysis of CoCs, this pilot study will provide critical insights into the distribution of potentially hazardous products within vulnerable communities.

By linking store census data with neighborhood-level demographic and socioeconomic indicators, Project CLEAR further situates its findings within broader patterns of structural inequity and environmental injustice. The chemical analyses, employing advanced non-targeted methods, will allow for the identification of both listed and undisclosed toxicants, offering a more comprehensive understanding of consumer risks.

The results of this pilot study will lay the foundation for larger investigations into the health impacts of SLP use, inform regulatory efforts, and support community-centered advocacy for safer consumer products. As reviewed and deemed exempt by the Columbia University Institutional Review Board (IRB), Project CLEAR represents an important step toward illuminating hidden drivers of environmental health disparities and promoting equity in consumer safety.

## Data Availability

No complete datasets were generated yet as pilot data was solely used to modify field research protocol. All relevant data from this study will be made available upon study completion.

## Acknowledgements

This study was partly supported by the NIEHS P30 Columbia Center for Environmental Health and Justice in Northern Manhattan (P30ES009089) and the Department of Epidemiology at the Mailman School of Public Health at Columbia University.

